# Smoking Cessation, Weight Change, and Risk of Dementia: A Prospective Cohort Study

**DOI:** 10.1101/2025.11.03.25339447

**Authors:** Hui Chen, Jingjing Wang, Sirui Lai, Guoping Peng, Geng Zong, Changzheng Yuan, Benyan Luo

## Abstract

**Objectives:** To assess the associations of smoking cessation and post-cessation weight gain with the risk of dementia and cognitive trajectories.

**Design:** Prospective cohort study.

**Setting:** The U.S. Health and Retirement Study (1995-2020).

**Participants:** A total of 32,802 dementia-free participants were included, with a mean age of 60.5 years (SD 10.7) and 57.1% female.

**Exposure:** Smoking status and body weight were collected biennially via structural interviews.

**Main outcome measures:** Dementia was identified using the Langa-Weir algorithm. Cognitive function was assessed using a 27-unit scale. Cox proportional hazard models estimated hazard ratio (HR) of dementia by smoking cessation status, subsequent weight change, and duration of cessation. Among participants who quit during follow-up, linear mixed models assessed cognitive trajectories before and after cessation.

**Results:** Over 25 years of follow-up, 5,868 dementia cases were documented. Compared with current smokers, individuals who quit during follow-up had a lower dementia risk after quitting (HR: 0.82, 95% confidence interval: 0.72-0.93), similar to those who had quit before baseline (0.76, 0.69-0.83) and to never smokers (0.72, 0.66-0.79). The benefits of cessation were largely limited to participants with no or modest weight gain (≤5 kg). By contrast, quitting accompanied by >10 kg weight gain was marginally associated with higher dementia risk (1.31, 0.95-1.80). Dementia risk declined steadily with increasing cessation duration, reaching the level of never smokers after approximately 5-7 years. Cognitive trajectory analyses showed that quitting was associated with long-term slower cognitive decline but no transient change, especially among those with no or modest weight gain.

**Conclusions:** Smoking cessation was associated with a sustained lower dementia risk and slower cognitive decline, comparable to benefits observed in never smokers and without evidence of a short-term risk increase. However, substantial post-cessation weight gain may attenuate these advantages. Smoking cessation programs should incorporate weight-management strategies to optimize long-term brain health.

**SUMMARY BOX:**

- What is already known on this topic
- Smoking cessation is universally prioritized for reducing cardiovascular and cancer risk.
- Weight gain commonly occurs after quitting and can lead to adverse metabolic outcomes.
- So far, few studies have examined how cessation timing, duration, and post-cessation weight changes influence long-term dementia risk.

**What this study adds:**

- In this study of 32,802 U.S. middle-aged and older adults, smoking cessation was associated with a steadily declining dementia risk, reaching the level of never smokers after approximately 5-7 years.
- Substantial post-cessation weight gain may attenuate these advantages, and the lower risk following cessation were largely limited to participants with no or modest weight gain (≤5 kg).
- Smoking cessation was also associated with long-term slower cognitive decline with no transient change, especially among those with no or modest weight gain.

## INTRODUCTION

Dementia is a leading cause of disability and dependence among older adults worldwide, and its global prevalence is expected to rise sharply in the coming decades.[1,2] Identifying modifiable risk factors is therefore a public health priority.[3,4] Smoking is an established contributor to vascular disease, oxidative stress, and neurodegeneration, as well as increase the risk of cerebrovascular disease and diabetes, both of which are established risk factors for dementia.[1] While numerous studies have suggested that current smokers are at higher risk of cognitive decline and dementia, evidence regarding the impact of smoking cessation on dementia risk has been rare and inconsistent.[5–7] In these studies, former smokers experience risk reductions approaching those of never smokers, but the effect size varied.[5,6]

A potential source of uncertainty emerged from the health consequences of weight gain following smoking cessation.[8,9] Weight gain is a common outcome after quitting, driven by increased appetite, reduced basal metabolic rate, and improvements in taste and olfactory sensitivity associated with nicotine withdrawal.[8,10] In a cohort study of ∼170,000 participants, recent quitters had a 22% higher risk of type 2 diabetes compared with current smokers, with the excess risk peaking within 5-7 years and diminishing thereafter.[11] Crucially, the adverse metabolic effects were more pronounced in individuals who gained weight.[11,12] Despite this short-term concern, the risk of mortality declined with cessation across most populations.[11,13,14] Given that both metabolic disorders and cardiovascular disease are recognized risk factors for dementia, these findings raise the question of whether post-cessation weight gain may modify the cognitive benefits of quitting. Current studies on smoking cessation and dementia are largely based on point prevalence abstinence (assessment of smoking status at baseline only),[5,15] which may underestimate the health consequences of smoking.[16] In addition, while successful smoking quitters have more favorable brain structural changes,[17] evidence on the role of post-cessation weight gain in shaping dementia risk is scarce, especially during a long-term follow-up.

To address these gaps, we analyzed data from the Health and Retirement Study (HRS), a nationally representative cohort of U.S. adults followed for over two decades. We examined the associations of smoking cessation and subsequent weight change with incident dementia and further investigated the trajectories of cognitive function after cessation. We hypothesized that smoking cessation would be associated with reduced dementia risk, but that excessive weight gain following cessation might attenuate or even offset these benefits.

## METHODS

### Study design and participants

We conducted a prospective cohort study using data from the Health and Retirement Study (HRS), a nationally representative longitudinal survey of U.S. adults aged 50 years and older.[18] The HRS collects detailed information on sociodemographic characteristics, health behaviors, medical history, and cognitive function through biennial interviews, with comprehensive cognitive and dietary assessments administered in selected waves. The design and sampling procedures of the HRS have been described in detail elsewhere.[18]

For the present analysis, we included participants from multiple HRS entry cohorts. We included the HRS original cohort (born 1931-1941, enrolled in 1992) and the AHEAD cohort (born before 1924, enrolled in 1993-1994) from the 1995-1996 wave, when cognitive assessments required for the Langa-Weir algorithm to define dementia first became available. Additional cohorts were incorporated at their respective baseline interviews, including the Children of the Depression (born 1924-1930) and War Babies (born 1942-1947) in 1998, Early Baby Boomers (born 1948-1953) in 2004, Mid Baby Boomers (born 1954-1959) in 2010, and Late Baby Boomers (born 1960-1965) in 2016. Participants were eligible if they had data on smoking status and were free of dementia at baseline. We excluded individuals with no follow-up interviews, as they did not contribute person-years to the analysis (**Figure S1**), and the baseline characteristics of included versus excluded participants were shown in **Table S1**. Person-years were calculated from the date of the baseline interview to the date of dementia diagnosis, death, loss to follow-up, or the end of the study period (2020), whichever occurred first.

### Assessment of smoking status and weight change

At each biennial interview, participants reported whether they had ever smoked and whether they were currently smoking. Based on these responses, we classified participants into three groups: current smokers, former smokers, and never smokers. During follow-up, individuals who reported smoking in the previous wave but not in the current wave were considered to have quit smoking.[11,13] In a previous meta-analysis, self-reports of smoking are generally accurate and reliable.[19] For these participants, we defined the onset of quitting as the beginning of the current wave and calculated the duration of smoking cessation until relapse, the occurrence of study outcomes, or the end of follow-up. Missing smoking information was imputed using responses from the immediately prior wave.

Body weight was self-reported at every biennial interview. The validity of self-reported weight has been previously documented.[20,21] For the analysis of weight change, we focused on two-year weight differences among participants who had recently quit smoking (**Figure S2**). This approach allowed us to capture weight changes from pre-to post-cessation without overlapping with subsequent follow-up, thereby reducing the potential for reverse causality.

### Assessment of cognitive function and dementia

Cognitive status in the HRS was assessed using a validated 27-point scale administered to self-respondents, including immediate and delayed recall (0-20 points), serial sevens subtraction (0-5 points), and counting backwards (0-2 points).[22] Based on the Langa-Weir classification, scores were categorized as normal cognition (12-27), cognitively impaired but not demented (CIND, 7-11), and dementia (0-6).[23] For participants unable to complete the cognitive battery, proxy interviews were used to generate a validated 11-point proxy scale combining informant memory ratings, instrumental activities of daily living, and interviewer assessments, which was similarly classified into normal, CIND, and dementia. Participants were defined as having dementia if either the self-report or proxy assessment indicated dementia. Details were described in the **Supplementary Text**.

### Covariates

In this study, analyses were adjusted for demographic, lifestyle, and health-related factors, consistent with previous research.[6,11,24] Age was grouped in 5-year intervals, and study wave was included to account for period effects. Additional demographic covariates comprised self-reported gender, marital status (married/partnered vs. not), highest educational attainment (below high school, GED, high school graduate, some college, or college and above), and race (White, Black, or Other). Lifestyle variables included physical activity (vigorous activity ≥3 times per week vs. less), body mass index (BMI; <25.0, 25.0-29.9, or ≥30.0 kg/m²), and alcohol consumption (non-drinker, ≤1 drink/day, 1-2 drinks/day, or >2 drinks/day). Major chronic conditions (hypertension, diabetes, heart disease, and stroke) were all self-reported. Further details are provided in the **Supplementary Text**. Because the missing rates of the covariates were <10% (**Table S2**), we used a separate category to impute the missing values.[11]

### Statistical analysis

Baseline characteristics of participants were summarized according to smoking status at study entry. Continuous variables were presented as means with standard deviations (SDs), and categorical variables as counts with percentages. We used Cox proportional hazard regression to estimate hazard ratios (HRs) and 95% confidence intervals (CIs) for the association between smoking cessation and the incidence of dementia. Smoking status was defined in three ways: (1) current, never, or former smoker at each wave; (2) current, never, quitters before baseline, and quitters during follow-up; and (3) quitters during follow-up further stratified by 2-year weight change categories (no weight gain, 0.1-5.0 kg, 5.1-10.0 kg, and >10.0 kg). Missing covariate values were handled using missing-indicator categories. We found no evidence of violation of the proportional hazard assumption for any exposure-outcome association via the Schoenfield’s residual methods. Models were stratified by age (in 5-year groups) and study wave and adjusted for gender, marital status, highest educational attainment, race, physical activity, BMI category, alcohol drinking status, and the prevalence of hypertension, diabetes, heart disease, and stroke.

To assess the association between the duration of smoking cessation and dementia, we fitted restricted cubic spline models with knots placed at standard percentiles of cessation duration. We additionally tested for interaction between cessation duration and 2-year weight change categories to examine whether post-cessation weight gain modified the association with dementia risk. To evaluate potential effect modification, we performed prespecified subgroup analyses by age (≥70 years vs. <70 years), sex, BMI category (<25 vs. ≥25 kg/m²), educational attainment (college or above vs. lower), and vigorous physical activity (≥3 times per week vs. less). Interaction terms between each subgroup variable and the primary exposure were tested using likelihood ratio tests.

For cognitive decline, we modeled cognitive trajectories using linear mixed-effects models with random intercepts and slopes to compare trajectories before and after smoking, and between quitters with ≥5.0 kg weight gain or <5.0 kg weight gain (**Supplementary Text**).

We conducted several sensitivity analyses to examine the robustness of the findings by: (1) excluding participants who reported major chronic diseases at baseline, including diabetes, heart disease, stroke, or cancer; (2) excluding participants developed dementia within the first two years after baseline; (3) excluding participants who were classified as cognitively impaired but not demented (CIND) at baseline; (4) excluding participants who developed dementia within the first two years after smoking cessation; and (5) performing competing risks analysis using Fine-Gray sub-distribution hazard model, treating death as a competing event for dementia incidence.

All P values were two-sided, with statistical significance defined as P < 0.05. Analyses were conducted using R software, version 4.5.0 (R Foundation for Statistical Computing).

### Patient and Public Involvement

Patients or the public were not involved in the design, or conduct, or reporting, or dissemination plans of our research.

## RESULTS

### Characteristics of the study participants

Among the 32,802 participants, the mean age at baseline was 60.5 years (SD 10.7) and 57.1% were female. The cohort was predominantly White (73.4%), with Black participants comprising 18.1% and other races 8.6%. Mean BMI in the whole sample was 27.9 kg/m² (SD 5.9). At study entry, 6,700 (20.4%) were current smokers, 11,951 (36.4%) were past smokers, and 14,151 (43.2%) had never smoked. Current smokers were younger (mean 57.2 years) than past smokers (62.2 years) and had lower educational attainment and household income. They are also less likely to have regular vigorous physical activity (32.9%). Alcohol drinking was more common and heavier among current and past smokers than among never smokers. (**Table 1**). Detailed categories of weight change among participants who quit smoking during follow-up are presented in **Table S3**.

**Table 1.**
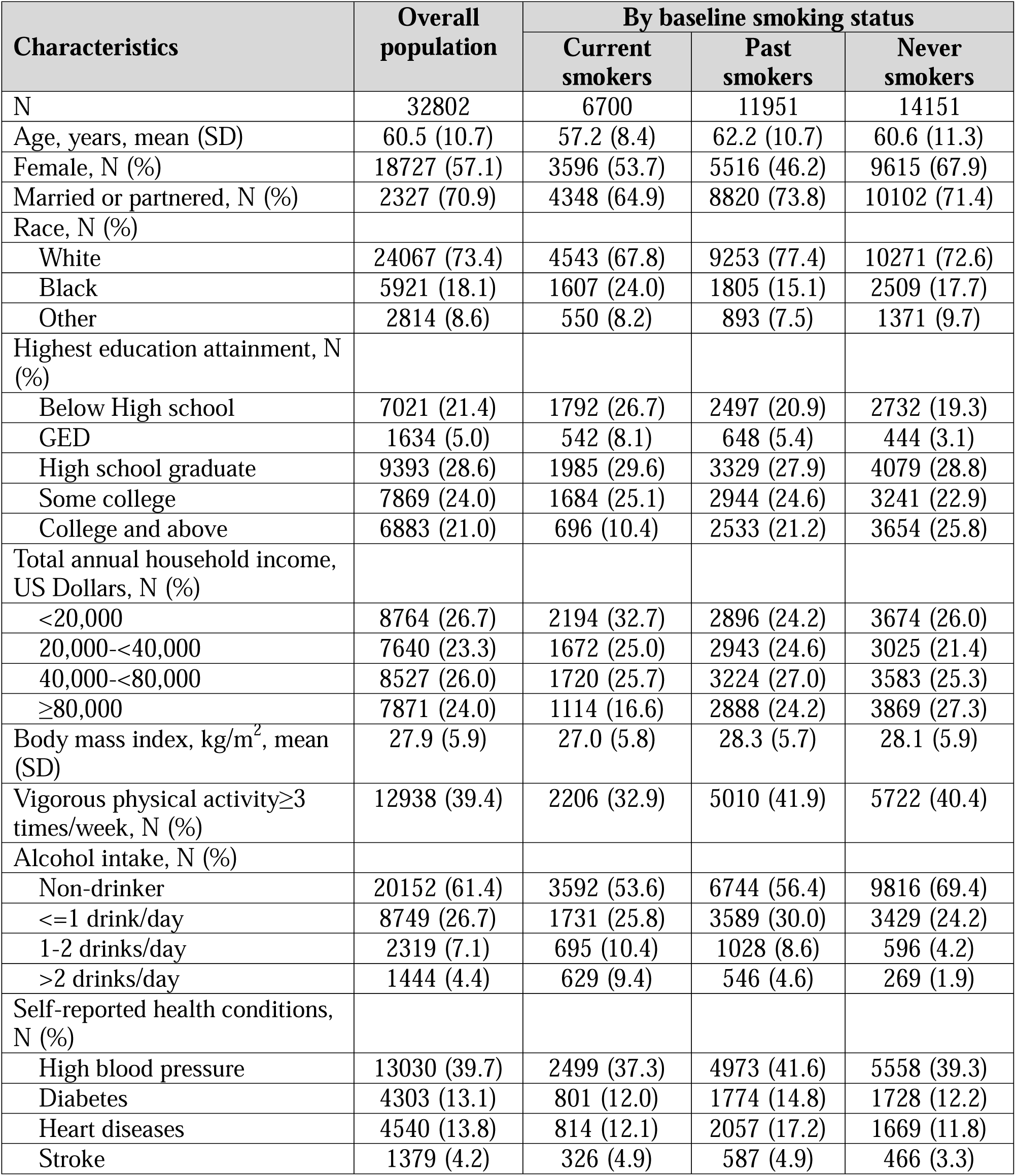
Characteristics of Study Participants at Baseline.

### Smoking cessation, weight change, and incident dementia

Over a maximum of 25 years of follow-up, we documented 5,868 incident dementia cases (**Table 2**). In age-and wave-stratified Cox models adjusted for sociodemographic factors, lifestyle and comorbidities (Model 3), smoking cessation was associated with a substantially lower risk of dementia compared with continued smoking. Compared with current smokers, past smokers (HR: 0.83, 95%CI: 0.76 to 0.90) and never smokers (0.78, 0.72 to 0.85) both had lower risk of dementia. The associations were congruent when we further separate those who had quit prior to baseline and during the study follow-up: HR 0.84 (0.73 to 0.95) for participants who quit during follow-up, and 0.79 (0.72 to 0.87) for those who had quit prior to baseline. The restricted cubic-spline analysis showed decreasing dementia risk with longer time since quitting, and the risk approached those of never smokers and plateaued at around 7 years post-cessation (**Figure 1A**).

**Figure 1.**
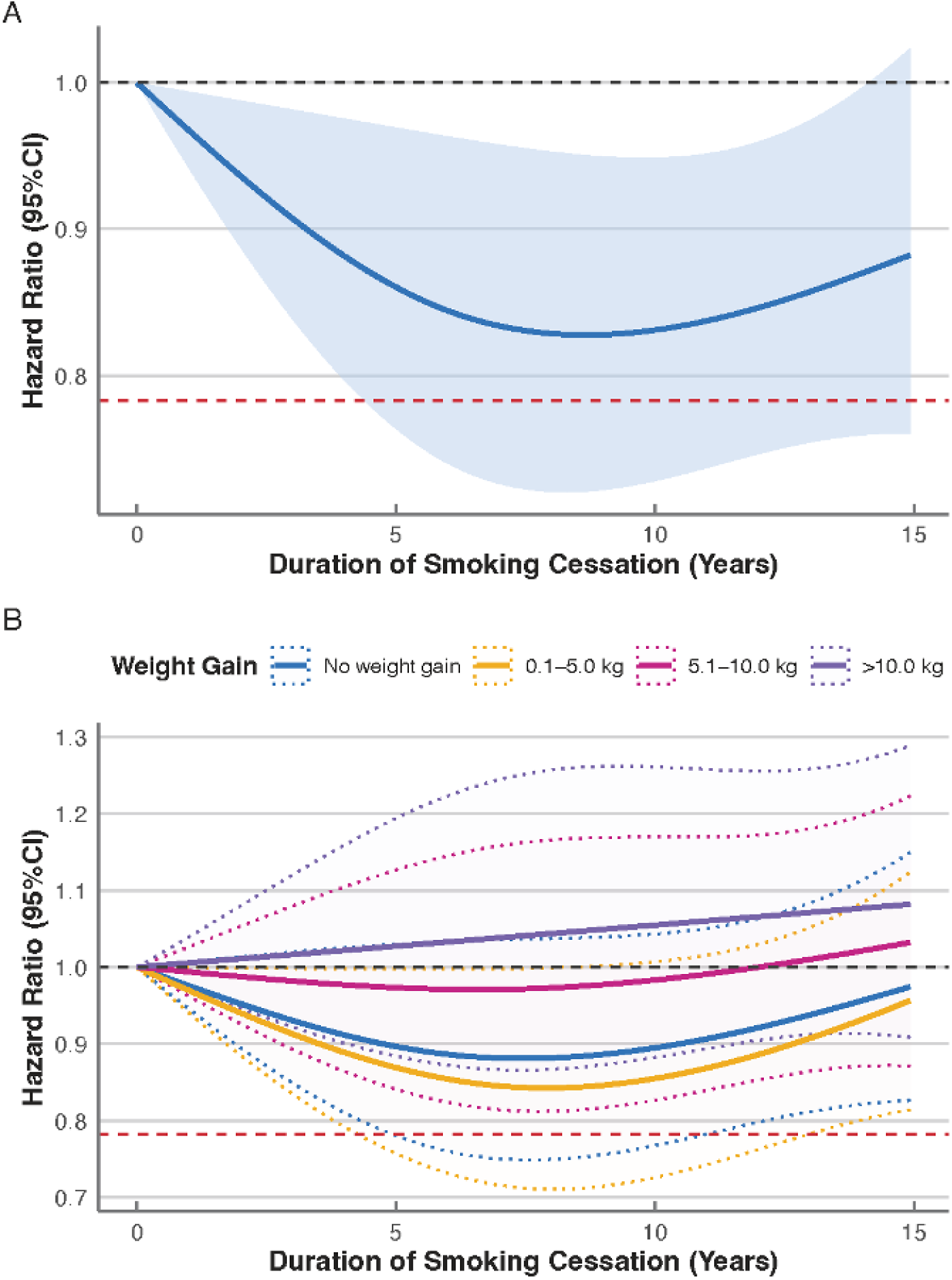
Restricted Cubic Splines for Association between Duration of Smoking Cessation and Risk of Dementia. The A panel shows the estimated hazard ratios and 95% CIs (shaded area) of dementia risk according to the duration of smoking cessation. The red dashed line indicated the hazard ratio for participants who never smoked, while the black dashed line indicated hazard ratio for current smokers or quitters (the reference group). The restricted cubic splines were fitted within a Cox proportional hazard model stratified by age (5-year groups) and study wave and adjusted for gender, marital status, highest education attainment, race, physical activity, BMI category, alcohol drinking status, and the prevalence of hypertension, diabetes, heart diseases, and stroke.

**Table 2.** Hazard Ratios of Dementia by Smoking Cessation Status.

| Variable | Cases/Person-<br>yrs | HR (95%CI) |  |  |
| --- | --- | --- | --- | --- |
|  |  | Model 1 | Model 2 | Model 3 |
| Concurrent smoking status |  |  |  |  |
| Current smokers | 812/54145 | 1 (Ref.) | 1 (Ref.) | 1 (Ref.) |
| Past smokers | 2448/152884 | **0.64 (0.59-0.70)** | **0.84 (0.77-0.92)** | **0.83 (0.76-0.90)** |
| Never smokers | 2608/162454 | **0.62 (0.57-0.67)** | **0.78 (0.72-0.85)** | **0.78 (0.72-0.85)** |
| Smoking quitting status |  |  |  |  |
| Current smokers | 702/46524 | 1 (Ref.) | 1 (Ref.) | 1 (Ref.) |
| Quitters, during follow-up | 358/22566 | **0.81 (0.71-0.93)** | **0.85 (0.74-0.97)** | **0.84 (0.73-0.95)** |
| Quitters, before baseline | 2198/137626 | **0.61 (0.55-0.66)** | **0.81 (0.74-0.89)** | **0.79 (0.72-0.87)** |
| Never smokers | 2610/162767 | **0.60 (0.55-0.66)** | **0.76 (0.69-0.83)** | **0.75 (0.69-0.83)** |
| Smoking quitting status and weight change |  |  |  |  |
| Current smokers | 702/46524 | 1 (Ref.) | 1 (Ref.) | 1 (Ref.) |
| Quitters, during follow-up |  |  |  |  |
| No weight gain | 154/9054 | 0.86 (0.72-1.03) | 0.84 (0.71-1.01) | **0.83 (0.69-0.99)** |
| Weight gain of 0.1-5.0 kg | 107/7846 | **0.68 (0.55-0.83)** | **0.74 (0.60-0.91)** | **0.73 (0.60-0.90)** |
| Weight gain of 5.1-10.0 kg | 55/3551 | 0.78 (0.59-1.03) | 0.86 (0.65-1.14) | 0.87 (0.66-1.15) |
| Weight gain of >10.0 kg | 42/2019 | 1.29 (0.94-1.77) | **1.40 (1.02-1.93)** | 1.33 (0.97-1.82) |
| Quitters, before baseline | 2198/137626 | **0.61 (0.55-0.66)** | **0.81 (0.74-0.89)** | **0.79 (0.72-0.87)** |
| Never smokers | 2610/162767 | **0.60 (0.55-0.66)** | **0.76 (0.69-0.83)** | **0.75 (0.69-0.83)** |

When quitters were stratified by post-cessation weight change, the cognitive benefits of quitting were confined to those with no or modest weight gain (*P*-value <0.001 for interaction). Compared with current smokers, quitters with no weight gain had an adjusted HR of 0.83 (0.69 to 0.99), and quitters who gained 0.1-5.0 kg had an HR of 0.73 (0.60 to 0.90). By contrast, quitters who gained 5.1-10.0 kg (0.87, 0.66 to 1.15) did not have a significantly lower risk, and those with the largest gains (>10.0 kg) showed a direction to higher risk (HR 1.33, 0.97 to 1.82), despite lack of statistical significance. In the restricted cubic-spline analysis, the risk of dementia was lower for quitters with no or 0.1-5.0 kg weight gain, but not among quitters who gained >5.0 kg (**Figure 1B**).

### Smoking cessation, weight change, and cognitive decline

Among the 3419 participants who quit during follow-up and who had repeated cognitive measurements, linear mixed-effects models showed no consistent abrupt improvement in cognition immediately at the time of cessation (point estimate: 0.57; 95% CI: -0.69 to 1.83) but a slower subsequent rate of cognitive decline compared with participants who continued smoking (**Table 3** and **Figure 2A**). Specifically, quitters experienced a change in the rate of decline of 0.19 points per decade (95% CI: 0.00 to 0.38) compared with continuing smokers, indicating a modest attenuation of decline after quitting. When stratified by post-cessation weight change (**Figure 2B**), the slowing of decline was present and statistically significant only among quitters with no or small weight gain of 0.1-5.0 kg gain (slope difference: 0.23 per decade, 95% CI 0.03 to 0.43). Quitters who gained >5.0 kg did not show a clear difference in slope compared with continuing smokers (change in slope 0.07, 95% CI -0.19 to 0.33).

**Figure 2.**
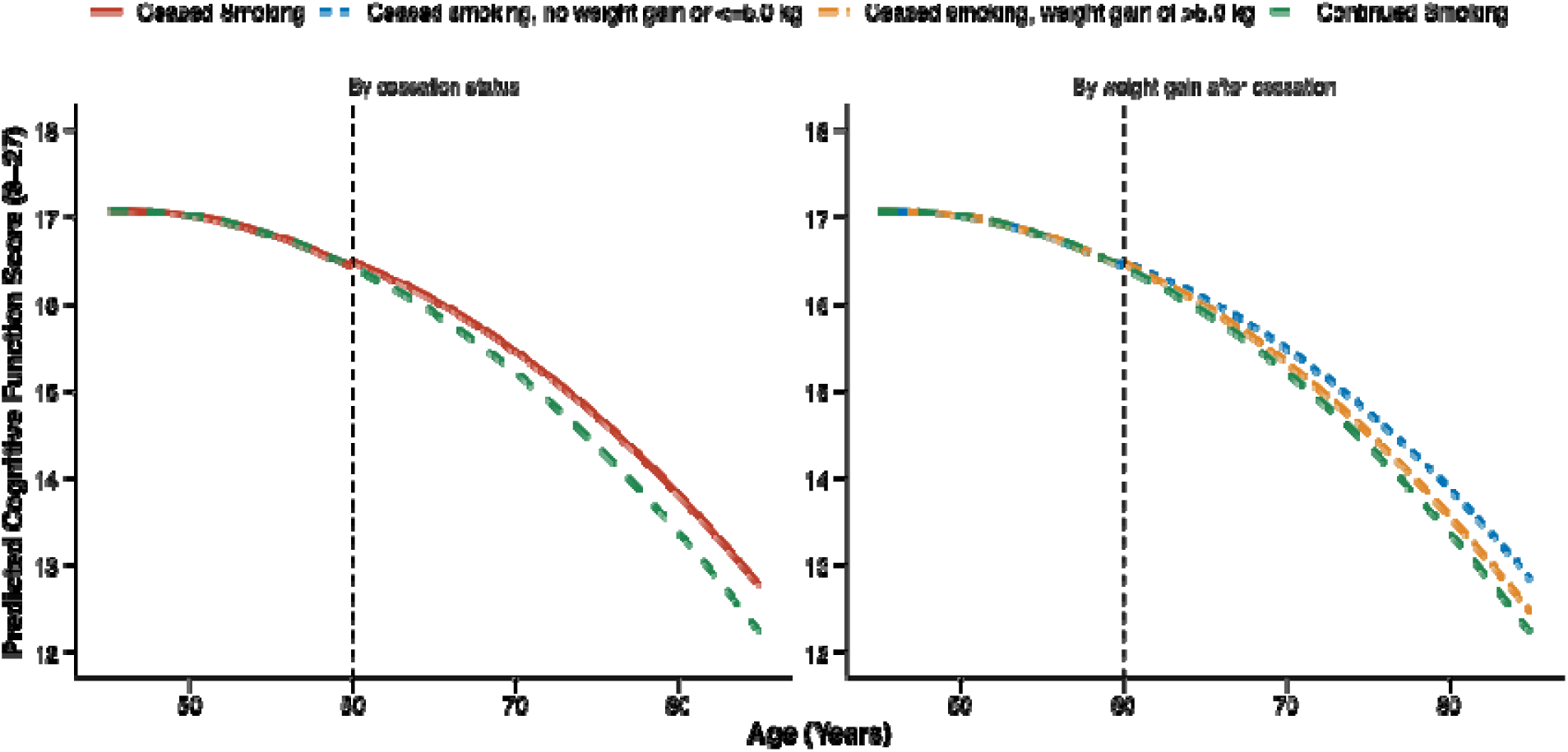
Fitted Trajectories of Cognitive Function by Smoking Cessation Status. The left panel shows the estimated cognitive trajectories and 95% CIs (shaded area) of a hypothetical person with smoking cessation at 60 years of age (population median age of cessation, presented as the vertical lines) in the reference categories of covariates (white race, female, married or partnered, 12 years of education, with vigorous physical activity >3 times/week, under or normal weight, no history of alcohol use, and no baseline comorbidities) compared with cognitive trajectories of a person with the same characteristics who remained smoking. The right panel further estimated the cognitive trajectories by weight gain around smoking cessation.

**Table 3.** Difference in Trajectories of Cognitive Change Per Decade Before and After Cessation Among Participants Quitting Smoking During Follow-up.

| Variable | Sudden Change in Cognitive Function | Change in Rate of Cognitive Decline with Linear Age |
| --- | --- | --- |
| By Smoking Status |  |  |
| Current Smokers | 0 (Ref.) | 0 (Ref.) |
| Quitters | 0.57 (-0.69, 1.83) | **0.19 (0.00, 0.38)** |
| By Weight Gain after Cessation |  |  |
| Current Smokers | 0 (Ref.) | 0 (Ref.) |
| Quitters with No or 0.1-5.0 kg Weight Gain | 0.54 (-0.82, 1.91) | **0.23 (0.03, 0.43)** |
| Quitters with Weight Gain of >5.0 kg | 0.44 (-1.67, 2.56) | 0.07 (-0.19, 0.33) |

Footnote: Estimates are from linear mixed-effects models with random intercepts, adjusted for demographic, lifestyle and health-related covariates. Sudden change reflects the immediate shift in cognition at cessation and change in rate of decline reflects the difference in slope of cognitive decline per decade compared with current smokers. Details of the models were described in Supplementary Text.

### Subgroup and sensitivity analyses

The association between smoke cessation with dementia were generally similar across study subgroups defined by age, BMI at baseline, and education attainment (**Table S4**). However, the association was stronger among men (HR: 0.70, 95%CI: 0.57 to 0.86) compared to women (0.96, 0.81 to 1.15, P-interaction = 0.05), and stronger among quitters engaging in regular physical activity (0.76, 0.60-0.96) that those who did not (0.91, 0.77 to 1.07). In terms of weight gain after cessation, quitters who gained between 0.1 and 5.0 kg showed the most significant reduction in dementia risk, while those who gained more than 10 kg had a higher dementia risk across most subgroups, particularly in older participants and those with higher BMI (**Table S5**).

The sensitivity analyses (**Table S6**) generally supported the robustness of the associations between smoking cessation and dementia risk. Excluding participants with major chronic diseases at baseline, those who developed dementia within the first two years, or participants with baseline cognitive impairment, yielded results consistent with the primary analysis. When we excluded participants who developed dementia within the first two years after smoking cessation, the associations were strengthened: the HR for quitters who had no weight gain during follow-up was 0.46 (95% CI: 0.36 to 0.58), stronger than that in the main analysis (0.83, 0.69 to 0.99). In addition, the competing risk analysis generated similar findings as in the primary analysis.

## DISCUSSION

In this prospective cohort study, smoking cessation was associated with a lower risk of dementia, with no short-term impact on cognitive function or elevated dementia risk. The greatest cognitive benefits were seen in individuals who experienced minimal to modest weight gain following cessation, while substantial post-cessation weight gain may attenuate these advantages. These findings highlight the importance of incorporating weight-management strategies in smoking cessation programs to optimize long-term brain health.

The association between smoking cessation and reduced dementia risk is consistent with previous studies suggesting that smoking is a major risk factor for cognitive decline and dementia. In a previous meta-analysis, current smokers have an 30% higher risk of all-cause dementia, 40% higher risk of AD, and 38% higher risk of VaD.[25] In a recent meta-analysis, smoking accounted for 4.5% of the incidence of dementia worldwide.[26] In our study, both former smokers and those who quit during the follow-up period had a significantly lower risk of dementia compared to current smokers, with the greatest benefit observed in those who quit smoking long-term. These findings align with the growing body of literature supporting the cognitive benefits of smoking cessation, particularly when cessation is sustained over time. In a cohort study in Japan (mean follow-up: 5.7 years), current smokers had a higher risk of incident dementia compared to never smokers, but ex-smokers who maintained abstinence for at least 3 years (retrospectively collected status) showed no increased risk compared with never smokers.[15] Similarly, in the Korean National Health Insurance System, long term quitters and never smokers at baseline had lower risk of dementia during a mean of 7.1 years.[27] More importantly, we did not observe a short-term elevated dementia risk in former smokers, which aligned with prior research that has documented short-term elevated risks for mortality, although diabetes risk increased in the first 5-7 years.[11]

The role of weight gain following smoking cessation in brain health remains largely unclear, and our study provides some novel insights. As noted in prior research, smoking cessation often leads to weight gain, driven by changes in appetite, metabolism, and sensory function.[8] While previous studies have highlighted the metabolic risks associated with post-cessation weight gain, particularly in relation to type 2 diabetes, our study extends this by examining the potential impact of weight gain on dementia risk. We found that post-cessation weight gain of 0.1-5.0 kg was associated with further reductions in dementia risk compared to continuing smoking. However, individuals who gained more than 5.0 kg experienced no significant cognitive benefit, and those with the highest weight gain (>10.0 kg) had a trend toward increased dementia risk, although this association did not reach statistical significance. This suggests that while the overall benefits of smoking cessation are clear, they do not necessarily outweigh the harms of substantial weight gain, and thus, excessive weight gain following smoking cessation may compromise the long-term cognitive advantages associated with quitting. Public health interventions should, therefore, not only encourage smoking cessation but also integrate weight management strategies to prevent excessive weight gain, which may negate some of the cognitive advantages of quitting smoking.

The relationship between weight gain and cognitive outcomes after smoking cessation is further supported by our analysis of cognitive decline trajectories. Quitters who gained little to no weight experienced slower rates of cognitive decline compared to continuing smokers. In contrast, those who gained significant weight (>5.0 kg) did not show an improvement in cognitive decline, reinforcing the notion that excessive weight gain may negate some of the cognitive benefits associated with smoking cessation. This finding underscores the need for tailored interventions that promote smoking cessation while mitigating the risk of excessive weight gain, which could otherwise undermine the long-term benefits to brain health.

The mechanisms by which smoking accelerates neurodegeneration have been widely studied, with evidence linking smoking to vascular damage, oxidative stress, and inflammation, all of which may contribute to cognitive decline and dementia.[28] This suggests that the benefits of smoking cessation on brain health may stem from the reversal or reduction of these harmful processes. In addition, smoking has been shown to exacerbate key pathological features of Alzheimer’s disease, such as amyloid plaque formation, neuroinflammation, and tau phosphorylation.[29,30] However, the potential adverse effects of excessive weight gain following smoking cessation, including increased inflammation and metabolic dysregulation, could complicate these benefits, highlighting the need for further research.[31] Future research should investigate the mechanisms through which weight gain influences brain health and explore potential interventions that could help individuals maintain a healthy weight after quitting smoking.

Strengths of this study include its large, nationally representative sample and extended follow-up period of up to 25 years, providing a robust basis for examining the long-term effects of smoking cessation on dementia risk. The repeated assessment of smoking status, weight change, and dementia outcomes, along with comprehensive adjustments for a wide range of potential confounding factors, allowed for a detailed analysis of smoking cessation trajectories during follow-up, the duration of smoking cessation, and post-cessation weight changes. Despite these strengths, several limitations should be noted. First, the use of self-reported smoking and weight data may introduce recall bias, though the large sample size and long follow-up period mitigate some of these concerns. Second, the study design is observational, so causality cannot be definitively established.

Although we have controlled for a range of potential confounders, unmeasured factors may still influence the observed associations. Additionally, the classification of smoking cessation based on self-reported smoking status may not capture the full complexity of smoking patterns, such as short-term intermittent smoking or relapse within the two-year intervals, which could affect the outcomes. Finally, although this study involves a wide range of races and ethnicities, whether the study findings may be generalized to other populations warrants investigation.

In conclusion, our findings emphasize the importance of smoking cessation for dementia prevention among smokers and that excessive weight gain following cessation may attenuate or even negate the cognitive benefits. These results suggest that public health interventions aimed at reducing smoking-related dementia risk should consider not only the promotion of smoking cessation but also weight management with quitting. Future research should further explore the mechanisms by which weight gain may influence brain health and investigate potential interventions to support both smoking cessation and weight management in aging populations.

## Data Availability

All data produced in the present study are available upon reasonable request to the authors

## Acknowledgements

We gratefully acknowledge Health and Retirement Study participants who contributed data for this study.

## Ethics approval

All Health and Retirement Study data collection protocols are approved by the University of Michigan Institutional Review Board.

## Funding

This work was supported by the supported by the Key R&D Program of Zhejiang (2022C03064, to BL). The funders had no role in considering the study design or in the collection, analysis, or interpretation of data, the writing of the report, or the decision to submit the article for publication.

## Competing interests

All authors have completed the ICMJE uniform disclosure form at www.icmje.org/disclosure-of-interest/ and declare: support from Key R&D Program of Zhejiang for the submitted work; no financial relationships with any organisations that might have an interest in the submitted work in the previous three years; no other relationships or activities that could appear to have influenced the submitted work.

## Contributors

HC, CY, and BL designed the study. HC, JW, SL, and GP contributed to the analysis or interpretation of the data. HC, JW, and SL prepared the first draft of the manuscript. All authors contributed to the interpretation of the results and to critical revision of the manuscript for important intellectual content and approved the final version of the manuscript. BL is the guarantor. The corresponding author attests that all listed authors meet authorship criteria and that no others meeting the criteria have been omitted.

## Transparency statement

The lead author (the manuscript’s guarantor) affirms that the manuscript is an honest, accurate, and transparent account of the study being reported; that no important aspects of the study have been omitted; and that any discrepancies from the study as planned (and, if relevant, registered) have been explained.

## Data availability statement

Data are available in a public, open access repository. The dataset used for this study was generated from data products publicly released by the Health and Retirement Study (HRS): https://hrs.isr.umich.edu. The HRS is sponsored by the National Institute on Ageing (grant number NIA U01AG009740).

## Dissemination to participants and related patient and public communities

The research findings will be disseminated to the public through press releases and conference presentations

## Notes

### Competing Interest Statement

The authors have declared no competing interest.

### Author Declarations

The study used ONLY openly available human data that were originally located at: https://hrs.isr.umich.edu

