## Supplement for "Smoking Cessation, Weight Change, and Risk of Dementia: A Prospective Cohort Study"

**Supplementary Materials**

### **Supplementary Methods**

#### Assessments of cognitive function and dementia

The HRS employs multiple measures to assess cognitive status. For self-respondents, we used the 27-point cognitive scale administered at each wave. This scale comprises: (1) immediate and delayed 10-word free recall tests to measure episodic memory (0–20 points); (2) a serial sevens subtraction task to assess working memory (0–5 points); and (3) a counting backwards test to measure processing speed (0–2 points). The Langa–Weir classification maps onto this scale as follows: normal cognition (12–27), cognitively impaired but not demented (CIND, 7–11), and dementia (0–6). Details of the classification were described previously.

To reduce sample attrition, HRS also incorporates proxy respondents when self-response is not possible. The proxy cognitive scale combines three measures: (1) proxy ratings of the respondent’s memory (excellent = 0, very good = 1, good = 2, fair = 3, poor = 4); (2) limitations in instrumental activities of daily living (0–5); and (3) interviewer assessments of cognitive impairment (no CI = 0, may have CI = 1, has CI = 2). This results in an 11-point scale, with classifications of normal (0–2), CIND (3–5), and dementia (6–11). In the 2000 interview, the interviewer assessment included a fourth category (“has CI”), which was coded as dementia. For waves prior to 2000 (1995–1998), the interviewer assessment of cognition was not administered, so a 9-point scale was constructed from the proxy memory ratings and IADL limitations (normal = 0–2, CIND = 3–4, dementia = 5–9).

For this study, we used the imputed information for the HRS cognitive measures. In all waves, participants were classified as having dementia if either the objective cognitive battery or the proxy scale indicated dementia.

#### Specifications of the mixed model for cognitive trajectory

To visualize the cognitive trajectory before and after smoking cessation, we specified a linear mixed-effects model assuming cessation at age 60, the median age of quitting in the cohort:

$$y_{\left\{ i,t \right\}}={\beta_{1}x}_{1}+{\beta_{2}x}_{2}+{\beta_{3}x}_{3}+{\beta_{4}x}_{4}+\beta_{5}x_{4}^{2}+\sum\beta_{k}x_{k}+\epsilon_{i}$$

In this framework, the binary variable $x_{1}$ indicated the exact timing of cessation, and therefore $\beta_{1}$ captured the immediate shift in cognitive score following cessation, and the continuous $x_{2}$ represent the linear time after cessation, allowing $\beta_{2}$ to represent the linear trend in cognitive change with age after cessation, with higher-order terms tested but excluded due to lack of significance. The continuous variable $x_{3}$ adjusted for heterogeneity in age at cessation by centering at 60 years (age at cessation - 60), and $x_{4}$ (age - 60) and $x_{4}^{2}$ flexibly modeled the age effect through linear and quadratic terms centered at 60. The model additionally adjusted for demographic and health-related covariates including gender, marital status, education, race, vaccination status, body mass index, alcohol use, hypertension, diabetes, heart disease, and stroke. A random intercept at the individual level accounted for within-person correlation across repeated measures. In analyses stratified by post-cessation weight gain, $\beta_{1}$ and $\beta_{2}$ were estimated for each category to allow heterogeneous trajectories across subgroups.

### **Supplementary Figures**

#### **Figure S1**. Study Population Inclusion Flow Chart

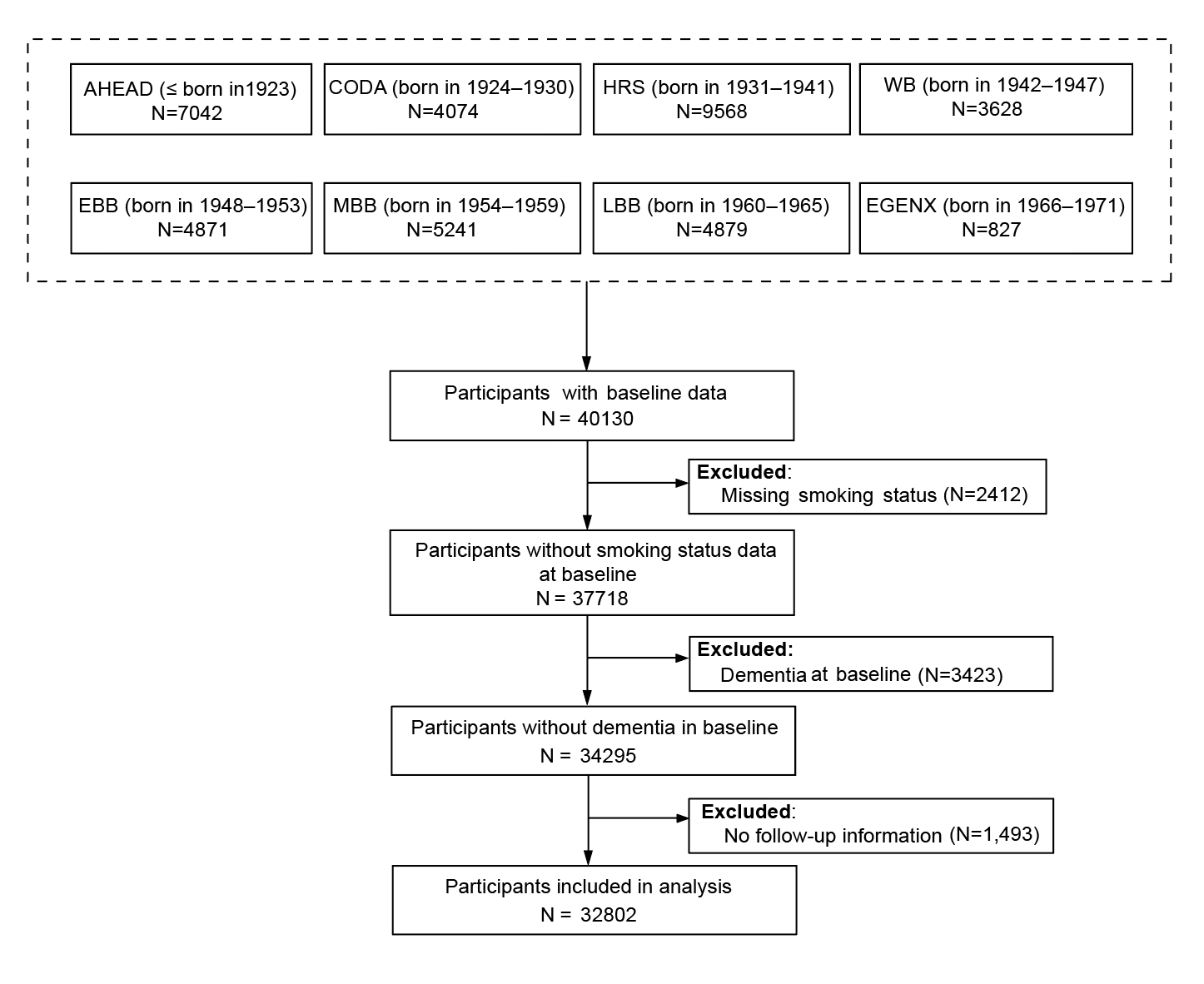

#### **Figure S2.** Mean Weight and Weight Change After Smoking Cessation Among Quitters During Follow-up

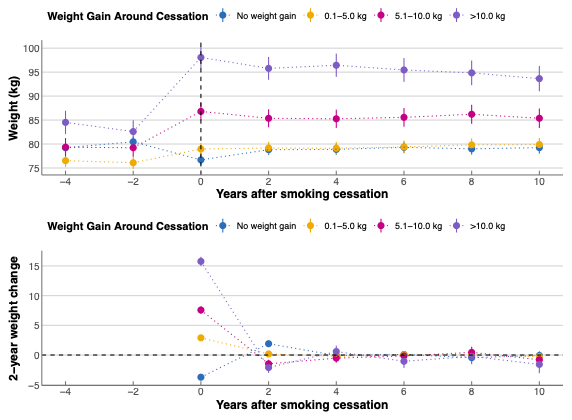

The least-square mean weights and weight changes across years after smoking cessation (as 2-year intervals) and weight change around cessation were estimated using linear mixed models including individual random intercept.

### **Supplementary Tables**

#### **Table S1**. Characteristics of Included vs. Excluded Participants at Baseline

| **Characteristics** | **Included**  **(n=32802)** | **Excluded**  **(n=7328)** | **Standardized Mean Difference** |
| --- | --- | --- | --- |
| Age, years, mean (SD) | 59.6 (10.3) | 65.6 (13.5) | 0.506 |
| Female, N (%) | 18727 (57.1) | 3904 (53.3) | 0.077 |
| Married or partnered, N (%) | 23270 (70.9) | 3285 (44.8) | 0.840 |
| Race, N (%) |  |  |  |
| White | 24067 (73.6) | 4827 (66.1) |  |
| Black | 5921 (18.1) | 1748 (23.9) |  |
| Other | 2732 (8.3) | 725 (9.9) |  |
| Highest education attainment, N (%) |  |  |  |
| Below High school | 7021 (21.4) | 2757 (37.7) |  |
| GED | 1634 (5.0) | 351 (4.8) |  |
| High school graduate | 9393 (28.6) | 1893 (25.9) |  |
| Some college | 7869 (24.0) | 1318 (18.0) |  |
| College and above | 6883 (21.0) | 993 (13.6) |  |
| Total annual household income, US Dollars, N (%) |  |  |  |
| <20,000 | 8764 (26.7) | 2566 (35.0) |  |
| 20,000-40,000 | 7640 (23.3) | 1200 (16.4) |  |
| 40,000-80,000 | 8527 (26.0) | 988 (13.5) |  |
| ≥80,000 | 7871 (24.0) | 782 (10.7) |  |
| Body mass index, kg/m^2^, mean (SD) | 27.9 (5.9) | 26.8 (6.1) | 0.203 |
| Vigorous physical activity ≥3 times/week, N (%) | 19835 (60.5) | 3993 (54.5) | 0.845 |
| Alcohol intake, N (%) |  |  |  |
| Non-drinker | 20152 (61.4) | 3918 (53.5) |  |
| ≤1 drink/day | 8749 (26.7) | 1061 (14.5) |  |
| 1-2 drinks/day | 2319 (7.1) | 287 (3.9) |  |
| >2 drinks/day | 1444 (4.4) | 225 (3.1) |  |
| Self-reported health conditions, N (%) |  |  |  |
| Diabetes | 4303 (13.1) | 1105 (20.0) | 0.207 |
| High blood pressure | 13030 (39.7) | 2767 (50.0) | 0.185 |
| Heart diseases | 4540 (13.8) | 1382 (25.0) | 0.284 |
| Stroke | 1379 (4.2) | 687 (12.4) | 0.301 |

#### **Table S2**. Missing Rates of Covariates

| **Characteristics** | **Missing Rates, %** |
| --- | --- |
| Age | 0 |
| Gender | 0 |
| Marital Status | 0.2 |
| Race | 8.6 |
| Highest education attainment | 0 |
| Total annual household income | 0 |
| Body mass index | 1.8 |
| Physical activity | 0.1 |
| Alcohol intake | 0.4 |
| Self-reported health conditions |  |
| High blood pressure | 0 |
| Diabetes | 0 |
| Heart diseases | 0 |
| Stroke | 0 |

#### **Table S3.** Characteristics of Study Participants Quitting Smoking During the Study Follow-up

| **Characteristics** | **All quitters** | **By weight gain after quitting** | | | |
| --- | --- | --- | --- | --- | --- |
|  |  | **No weight gain** | **0.1-5.0 kg** | **5.1-10.0 kg** | **>10.0 kg** |
| N | 2613 | 1131 | 839 | 394 | 249 |
| Age, years, mean (SD) | 62.2(8.6) | 62.5 (8.8) | 62.4 (8.3) | 61.5 (8.6) | 61.2 (8.2) |
| Female, N (%) | 1447 (55.4) | 628 (55.5) | 471 (56.1) | 210 (53.3) | 138 (55.4) |
| Married or partnered, N (%) | 1610 (61.6) | 662 (58.5) | 543 (64.7) | 254 (64.5) | 151 (60.6) |
| Race, N (%) |  |  |  |  |  |
| White | 1863 (71.3) | 754 (66.7) | 624 (74.4) | 301 (76.4) | 184 (73.9) |
| Black | 565 (21.6) | 278 (24.6) | 156 (18.6) | 73 (18.5) | 58 (23.3) |
| Other | 185 (7.1) | 99 (8.8) | 59 (7.0) | 20 (5.1) | 7 (2.8) |
| Highest education attainment, N (%) |  |  |  |  |  |
| Below High school | 612 (23.4) | 270 (23.9) | 188 (22.4) | 95 (24.1) | 59 (23.7) |
| GED | 178 (6.8) | 65 (5.7) | 66 (7.9) | 27 (6.9) | 20 (8.0) |
| High school graduate | 815 (31.2) | 355 (31.4) | 260 (31.0) | 112 (28.4) | 88 (35.3) |
| Some college | 694 (26.6) | 313 (27.7) | 206 (24.6) | 112 (28.4) | 63 (25.3) |
| College and above | 314 (12.0) | 128 (11.3) | 119 (14.2) | 48 (12.2) | 19 (7.6) |
| Total annual household income, US Dollars, N (%) |  |  |  |  |  |
| <20,000 | 828 (31.7) | 380 (33.6) | 241 (28.7) | 116 (29.4) | 91 (36.5) |
| 20,000-40,000 | 628 (24.0) | 290 (25.6) | 196 (23.4) | 92 (23.4) | 50 (20.1) |
| 40,000-80,000 | 677 (25.9) | 274 (24.2) | 235 (28.0) | 107 (27.2) | 61 (24.5) |
| ≥80,000 | 480 (18.4) | 187 (16.5) | 167 (19.9) | 79 (20.1) | 47 (18.9) |
| Body mass index before smoke cessation, kg/m^2^, mean (SD) | 27.5 (5.8) | 28.0 (6.0) | 26.5 (5.0) | 27.5 (5.9) | 28.3 (6.6) |
| Vigorous physical activity≥3 times/week, N (%) | 1919 (73.4) | 829 (73.3) | 585 (69.7) | 306 (77.7) | 199 (79.9) |
| Alcohol intake, N (%) |  |  |  |  |  |
| Never | 1709 (65.4) | 753 (66.6) | 532 (63.4) | 245 (62.2) | 179 (71.9) |
| ≤1 drink/day | 591 (22.6) | 262 (23.2) | 191 (22.8) | 99 (25.1) | 39 (15.7) |
| 1-2 drinks/day | 197 (7.5) | 78 (6.9) | 72 (8.6) | 30 (7.6) | 17 (6.8) |
| >2 drinks/day | 106 (4.1) | 34 (3.0) | 39 (4.6) | 19 (4.8) | 14 (5.6) |
| Self-reported health conditions, N (%) |  |  |  |  |  |
| High blood pressure | 1359 (52.0) | 595 (52.6) | 408 (48.6) | 199 (50.5) | 157 (63.1) |
| Diabetes | 476 (18.2) | 235 (20.8) | 135 (16.1) | 57 (14.5) | 49 (19.7) |
| Heart diseases | 593 (22.7) | 263 (23.3) | 181 (21.6) | 79 (20.1) | 70 (28.1) |
| Stroke | 217 (8.3) | 104 (9.2) | 64 (7.6) | 26 (6.6) | 23 (9.2) |

#### **Table S4**. Hazard Ratios of Dementia by Smoking Cessation Status (Quitters vs. Current Smokers) in Study Subgroups

| **Subgroup** | **Exposure** | **Cases/Person-yrs** | **HR (95% CI)** | **P for interaction** |
| --- | --- | --- | --- | --- |
| Age, years |  |  |  | 0.28 |
| <70 | Current smokers | 571 / 43406 | 1 (reference) |  |
| <70 | Quitters | 297 / 21216 | **0.82 (0.71–0.95)** |  |
| ≥70 | Current smokers | 131 / 3118 | 1 (reference) |  |
| ≥70 | Quitters | 61 / 1350 | 0.88 (0.64–1.21) |  |
| Sex |  |  |  | 0.05 |
| Male | Current smokers | 357 / 20240 | 1 (reference) |  |
| Male | Quitters | 149 / 9843 | **0.70 (0.57–0.86)** |  |
| Female | Current smokers | 345 / 26284 | 1 (reference) |  |
| Female | Quitters | 209 / 12723 | 0.96 (0.81–1.15) |  |
| BMI, kg/m^2^ |  |  |  | 0.12 |
| <25 | Current smokers | 336 / 18778 | 1 (reference) |  |
| <25 | Quitters | 128 / 8498 | **0.76 (0.61–0.94)** |  |
| ≥25 | Current smokers | 360 / 27347 | 1 (reference) |  |
| ≥25 | Quitters | 226 / 13782 | 0.88 (0.74–1.04) |  |
| Education |  |  |  | 0.30 |
| College and above | Current smokers | 38 / 4939 | 1 (reference) |  |
| College and above | Quitters | 14 / 2961 | **0.45 (0.24–0.86)** |  |
| Below college | Current smokers | 664 / 41574 | 1 (reference) |  |
| Below college | Quitters | 344 / 19605 | 0.88 (0.76–1.00) |  |
| Vigorous physical activity ≥3 times/week |  |  |  | 0.04 |
| No | Current smokers | 496 / 29331 | 1 (reference) |  |
| No | Quitters | 231 / 13099 | 0.91 (0.77–1.07) |  |
| Yes | Current smokers | 206 / 17173 | 1 (reference) |  |
| Yes | Quitters | 127 / 9464 | **0.76 (0.60–0.96)** |  |

Hazard ratios (HRs) and 95% confidence intervals (CIs) were estimated using Cox proportional-hazards models. Age (in 5-year groups) and study wave were included as stratification factors to account for age and period effects. For each subgroup analysis, the subgroup variable under investigation was not included as a covariate; all other covariates from the primary multivariable model were retained, including sex, marital status, education, race, physical activity, baseline body-mass index, alcohol intake, total household income, hypertension, diabetes, heart disease, and stroke. Tests for interaction were conducted by including a multiplicative term between the exposure and the subgroup variable in the model.

#### **Table S5.** Hazard Ratios of Dementia by Smoking Cessation Status (Quitters with Different Weight Gains vs. Current Smokers) in Study Subgroups

| **Subgroup** | **Exposure** | **Cases/Person-yrs** | **HR (95% CI)** | **P for interaction** |
| --- | --- | --- | --- | --- |
| Age, years |  |  |  | 0.05 |
| <70 | Current smokers | 571 / 43406 | 1 (reference) |  |
|  | Quitters |  |  |  |
|  | No weight gain | 132 / 8451 | 0.85 (0.70–1.03) |  |
|  | Weight gain of 0.1-5.0 kg | 89 / 7345 | **0.73 (0.58–0.92)** |  |
|  | Weight gain of 5.1-10.0 kg | 42 / 3367 | 0.82 (0.60–1.13) |  |
|  | Weight gain of >10.0 kg | 34 / 1965 | 1.11 (0.78–1.59) |  |
| ≥70 | Current smokers | 131 / 3118 | 1 (reference) |  |
|  | Quitters |  |  |  |
|  | No weight gain | 22 / 603 | 0.69 (0.43–1.10) |  |
|  | Weight gain of 0.1-5.0 kg | 18 / 501 | 0.74 (0.44–1.23) |  |
|  | Weight gain of 5.1-10.0 kg | 13 / 184 | 1.17 (0.66–2.09) |  |
|  | Weight gain of >10.0 kg | 8 / 54 | **3.24 (1.57–6.69)** |  |
| Sex |  |  |  | 0.20 |
| Male | Current smokers | 357 / 20240 | 1 (reference) |  |
|  | Quitters |  |  |  |
|  | No weight gain | 65 / 3902 | **0.65 (0.50–0.86)** |  |
|  | Weight gain of 0.1-5.0 kg | 43 / 3342 | **0.63 (0.46–0.87)** |  |
|  | Weight gain of 5.1-10.0 kg | 25 / 1680 | 0.83 (0.55–1.26) |  |
|  | Weight gain of >10.0 kg | 16 / 903 | 1.10 (0.66–1.84) |  |
| Female | Current smokers | 345 / 26284 | 1 (reference) |  |
|  | Quitters |  |  |  |
|  | No weight gain | 89 / 5152 | 1.01 (0.79–1.28) |  |
|  | Weight gain of 0.1-5.0 kg | 64 / 4504 | 0.82 (0.62–1.08) |  |
|  | Weight gain of 5.1-10.0 kg | 30 / 1871 | 0.94 (0.64–1.37) |  |
|  | Weight gain of >10.0 kg | 26 / 1116 | **1.52 (1.00–2.29)** |  |
| BMI, kg/m^2^ |  |  |  | 0.31 |
| <25 | Current smokers | 336 / 18778 | 1 (reference) |  |
|  | Quitters |  |  |  |
|  | No weight gain | 57 / 3305 | 0.83 (0.62–1.11) |  |
|  | Weight gain of 0.1-5.0 kg | 39 / 3306 | **0.62 (0.44–0.88)** |  |
|  | Weight gain of 5.1-10.0 kg | 21 / 1303 | 0.82 (0.52–1.28) |  |
|  | Weight gain of >10.0 kg | 11 / 584 | 0.93 (0.50–1.71) |  |
| ≥25 | Current smokers | 360 / 27347 | 1 (reference) |  |
|  | Quitters |  |  |  |
|  | No weight gain | 96 / 5663 | 0.86 (0.68–1.09) |  |
|  | Weight gain of 0.1-5.0 kg | 67 / 4483 | 0.78 (0.59–1.02) |  |
|  | Weight gain of 5.1-10.0 kg | 34 / 2236 | 0.84 (0.58–1.19) |  |
|  | Weight gain of >10.0 kg | 29 / 1400 | **1.47 (1.00–2.15)** |  |
| Education |  |  |  | 0.58 |
| College and above | Current smokers | 38 / 4939 | 1 (reference) |  |
|  | Quitters |  |  |  |
|  | No weight gain | 4 / 1138 | 0.40 (0.14–1.14) |  |
|  | Weight gain of 0.1-5.0 kg | 6 / 1154 | **0.38 (0.15–0.92)** |  |
|  | Weight gain of 5.1-10.0 kg | 2 / 497 | 0.52 (0.12–2.22) |  |
|  | Weight gain of >10.0 kg | 2 / 151 | 2.00 (0.46–8.66) |  |
| Below college | Current smokers | 664 / 41574 | 1 (reference) |  |
|  | Quitters |  |  |  |
|  | No weight gain | 150 / 7916 | 0.88 (0.73–1.05) |  |
|  | Weight gain of 0.1-5.0 kg | 101 / 6693 | **0.77 (0.62–0.95)** |  |
|  | Weight gain of 5.1-10.0 kg | 53 / 3054 | 0.91 (0.68–1.20) |  |
|  | Weight gain of >10.0 kg | 40 / 1868 | 1.31 (0.94–1.81) |  |
| Vigorous physical activity ≥3 times/week |  |  |  | 0.05 |
| No | Current smokers | 496 / 29331 | 1 (reference) |  |
|  | Quitters |  |  |  |
|  | No weight gain | 109 / 5504 | 0.92 (0.75–1.15) |  |
|  | Weight gain of 0.1-5.0 kg | 52 / 4261 | **0.70 (0.52–0.94)** |  |
|  | Weight gain of 5.1-10.0 kg | 41 / 2031 | 1.06 (0.76–1.46) |  |
|  | Weight gain of >10.0 kg | 29 / 1219 | 1.43 (0.97–2.10) |  |
| Yes | Current smokers | 206 / 17173 | 1 (reference) |  |
|  | Quitters |  |  |  |
|  | No weight gain | 45 / 3550 | **0.69 (0.50–0.97)** |  |
|  | Weight gain of 0.1-5.0 kg | 55 / 3584 | 0.79 (0.58–1.07) |  |
|  | Weight gain of 5.1-10.0 kg | 14 / 1520 | 0.61 (0.35–1.06) |  |
|  | Weight gain of >10.0 kg | 13 / 801 | 1.37 (0.78–2.43) |  |

Hazard ratios (HRs) and 95% confidence intervals (CIs) were estimated using Cox proportional-hazards models. Age (in 5-year groups) and study wave were included as stratification factors to account for age and period effects. For each subgroup analysis, the subgroup variable under investigation was not included as a covariate; all other covariates from the primary multivariable model were retained, including sex, marital status, education, race, physical activity, baseline body-mass index, alcohol intake, total household income, hypertension, diabetes, heart disease, and stroke. Tests for interaction were conducted by including a multiplicative term between the exposure and the subgroup variable in the model.

#### **Table S6.** Hazard Ratios of Dementia by Smoking Cessation Status in Sensitivity Analyses

| **Variable** | **Primary analysis** | **Sensitivity Analysis** | | | | |
| --- | --- | --- | --- | --- | --- | --- |
|  |  | **Sensitivity Analysis 1** | **Sensitivity Analysis 2** | **Sensitivity Analysis 3** | **Sensitivity Analysis 4** | **Sensitivity Analysis 5** |
| Concurrent smoking status |  |  |  |  |  |  |
| Current smokers | 1 (reference) | 1 (reference) | 1 (reference) | 1 (reference) | 1 (reference) | 1 (reference) |
| Past smokers | **0.83 (0.76-0.90)** | **0.83 (0.75-0.93)** | **0.83 (0.76-0.91)** | **0.86 (0.77-0.96)** | **0.74 (0.68-0.81)** | **0.88 (0.79-0.96)** |
| Never smokers | **0.78 (0.72-0.85)** | **0.80 (0.72-0.89)** | **0.77 (0.70-0.85)** | **0.78 (0.70-0.88)** | **0.76 (0.69-0.82)** | **0.89 (0.80-0.97)** |
| Smoking quitting status |  |  |  |  |  |  |
| Current smokers | 1 (reference) | 1 (reference) | 1 (reference) | 1 (reference) | 1 (reference) | 1 (reference) |
| Quitters, during follow-up | **0.84 (0.73-0.95)** | **0.84 (0.71-0.98)** | **0.86 (0.75-0.99)** | 0.89 (0.76-1.05) | **0.51 (0.44-0.59)** | **0.78 (0.65-0.91)** |
| Quitters, before baseline | **0.79 (0.72-0.87)** | **0.79 (0.71-0.89)** | **0.79 (0.72-0.88)** | **0.81 (0.72-0.91)** | **0.72 (0.66-0.79)** | **0.84 (0.75-0.93)** |
| Never smokers | **0.75 (0.69-0.83)** | **0.77 (0.68-0.86)** | **0.75 (0.68-0.82)** | **0.75 (0.66-0.84)** | **0.71 (0.65-0.78)** | **0.84 (0.75-0.93)** |
| Smoking quitting status and weight change |  |  |  |  |  |  |
| Current smokers | 1 (reference) | 1 (reference) | 1 (reference) | 1 (reference) | 1 (reference) | 1 (reference) |
| Quitters, during follow-up |  |  |  |  |  |  |
| No weight gain | **0.83 (0.69-0.99)** | 0.83 (0.67-1.03) | 0.86 (0.72-1.03) | 0.86 (0.69-1.07) | **0.46 (0.36-0.58)** | **0.78 (0.61-0.96)** |
| Weight gain of 0.1-5.0 kg | **0.73 (0.60-0.90)** | **0.78 (0.61-0.99)** | **0.75 (0.61-0.93)** | **0.77 (0.60-0.99)** | **0.48 (0.37-0.61)** | **0.68 (0.48-0.89)** |
| Weight gain of 5.1-10.0 kg | 0.87 (0.66-1.15) | 0.82 (0.58-1.15) | 0.88 (0.67-1.17) | 0.95 (0.69-1.29) | **0.60 (0.43-0.83)** | 0.81 (0.53-1.08) |
| Weight gain of >10.0 kg | 1.33 (0.97-1.82) | 1.33 (0.89-2.00) | 1.37 (0.99-1.88) | **1.55 (1.08-2.20)** | 0.78 (0.53-1.17) | 1.27 (0.96-1.59) |
| Quitters, before baseline | **0.79 (0.72-0.87)** | **0.79 (0.71-0.89)** | **0.79 (0.72-0.88)** | **0.81 (0.72-0.91)** | **0.72 (0.66-0.79)** | **0.84 (0.75-0.93)** |
| Never smokers | **0.75 (0.69-0.83)** | **0.77 (0.68-0.86)** | **0.75 (0.68-0.82)** | **0.75 (0.66-0.84)** | **0.71 (0.65-0.78)** | **0.84 (0.75-0.93)** |

Four sensitivity analyses were conducted to evaluate the robustness of the findings. (1) Sensitivity analysis 1 excluded participants who reported major chronic diseases at baseline, including diabetes, heart disease, stroke, or cancer (*n* = 22,809). (2) Sensitivity analysis 2 excluded individuals who developed dementia within the first two years after baseline, to minimize reverse causality (*n* = 32,388). (3) Sensitivity analysis 3 excluded participants with cognitively impaired but not demented (CIND) at baseline (*n* = 28,619). (4) Sensitivity analysis 4 excluded participants who developed dementia within the first two years after smoking cessation (*n* = 32,584). (5) Sensitivity analysis 5 was performed using the Fine-Gray sub-distribution hazard model, treating death as a competing event for dementia incidence.
